# Empowering Self-Management for Chronic Low Back Pain: A Human-Centered Design Study of Spanish- and Cantonese-Preferring Patients in the United States

**DOI:** 10.1101/2024.09.27.24314504

**Authors:** Patricia Zheng, Emilia De Marchis, Jan Yeager, Karina Del Rosario, Masato Nagao, Tigist Belaye, Angela Gallegos-Castillo, Lei-Chun Fung, Adrian Vallejo, Amy Kuang, David Gendelberg, Jeffrey Lotz, Conor O’Neill, the REACH investigators

## Abstract

**Introduction:** Chronic low back pain (cLBP) is a leading cause of disability with disproportionately high impacts on marginalized populations, including non-English-preferring patients. These patients face significant barriers to accessing care and adhering to self-management strategies due to language barriers, socioeconomic challenges, and cultural differences. Despite the emphasis on self-management for cLBP, limited research has focused on understanding the specific needs and preferences of Spanish- and Cantonese-preferring patients.

**Objective:** This study aimed to explore the self-management priorities of Spanish- and Cantonese-preferring patients with cLBP. Using a human-centered design approach, we sought to identify patient preferences for self-management support materials and strategies that could be tailored to their unique needs.

**Design:** Qualitative research using thematic analysis of focus groups conducted in participants’ preferred language.

**Setting:** Urban, academic-affiliated county hospital between March and May 2024.

**Patients:** Spanish- and Cantonese-preferring patients with cLBP

**Interventions:** Not applicable.

**Main outcome:** Key themes in participants’ experiences with cLBP care, barriers to self-management, and preferences for educational materials.

**Results:** Fifteen patients participated across six focus groups (three focus group in each language). Four primary themes emerged from the focus groups: (1) the need for empathic, tailored educational supports that fit into patients’ lives, (2) a desire for self-management plans that account for social and economic constraints, (3) recognition of mental health and social isolation as factors that influence cLBP experience, and (4) a need for clearer guidance on self-management strategies and trustworthy resources. Both Spanish- and Cantonese-preferring participants expressed a preference for video-based resources, plain-language materials, and support for understanding the causes and management of their pain.

**Conclusion:** Spanish- and Cantonese-preferring patients with cLBP face significant barriers to self-management and would benefit from culturally and linguistically appropriate resources. This study highlights the need for healthcare systems to develop and deliver tailored, accessible self-management support materials that address the unique challenges faced by minoritized populations. Human-centered design offers a promising approach to reducing disparities in cLBP outcomes by creating patient-driven solutions that prioritize empathy, practicality, and cultural relevance.

## Introduction

Chronic low back pain (cLBP) is one of the leading causes of disability worldwide, affecting a diverse range of populations across different socioeconomic and cultural contexts.^1–3^ The burden of cLBP is particularly high among marginalized groups, including those who face language, racial, and cultural barriers to healthcare.^4–19^ Language preference, often acting as a proxy for broader issues like racism and discrimination, plays a critical role in shaping access to care, treatment outcomes, and patient satisfaction.^20–22^ For patients who prefer to speak non-English-languages in the United States, such as Spanish- and Cantonese-speaking populations, these barriers are further compounded by socioeconomic factors such as poverty, limited education, and restricted access to resources.^23,24^ These structural inequities contribute to the disproportionately high levels of cLBP-related disability and dissatisfaction with care in these communities.^19,25^

Current approaches to cLBP management emphasize patient engagement through self-management strategies, which include physical therapy, exercise, lifestyle changes, and psychological support.^26–28^ However, existing self-management programs often fail to account for the unique challenges faced by non-English-speaking patients.^29,30^ These patients may have difficulty understanding care instructions, lack access to culturally and linguistically appropriate educational materials, and face socioeconomic barriers that make it difficult to prioritize self-care.^24,30,31^ Moreover, much of the existing literature on cLBP care overlooks the specific needs of non-English-preferring populations, particularly in the context of low-income, urban environments.^30^ This gap in research underscores the need for a more nuanced understanding of how to support these patients in managing their cLBP.

Human-centered design (HCD) offers a promising approach to addressing disparities in cLBP outcomes and increasing access to self-management resources by focusing on the lived experience of patients and developing interventions that are tailored to their specific needs.^32^ Unlike traditional top-down healthcare models, HCD engages patients as co-creators in the design process, ensuring that solutions are not only culturally and linguistically appropriate but also practical and feasible within the constraints of their daily lives. By placing patients at the center of the design process, HCD can help identify the priorities and preferences that matter most to them, particularly in relation to self-management education.^33,34^

This qualitative study aimed to explore the cLBP self-management priorities of Spanish- and Cantonese-preferring patients at an urban, academic-affiliated county hospital. Using a HCD approach, we sought to understand how existing self-management resources could be optimized to better meet the needs of these populations.

## Methods

### Study Design and Setting

This study was approved by the Institutional Review Board (IRB) at the University of California, San Francisco and took place at an urban, academic-affiliated county hospital. All participants provided written informed consent prior to their involvement in the study. We adhered to the Consolidated Criteria for Reporting Qualitative Research (COREQ) guidelines throughout the study design, data collection, and analysis processes (see Appendix X for COREQ checklist).

### Participant Recruitment

We recruited a convenience sample of adult patients seen in a clinic visit between January and May 2024. Patients were eligible for participation if they met the NIH Research Task Force criteria^35^ for chronic low back pain: current self-report of chronic low back pain (pain between the lower posterior margin of the rib cage and the horizontal gluteal fold), which has persisted for at least the past 3 months AND has resulted in pain on >50% of days in the past 6 months. Participants were included if they reported preferring to speak either Spanish or Cantonese during healthcare interactions. We identified patients via chart review and clinician referrals, and those who met the inclusion criteria were contacted by phone or in person in their preferred language by bilingual study team members or using a certified phone interpreter to ensure accurateness of interpretation.

Eligible patients were invited to participate in focus groups and informed that participation was voluntary, would not impact their care, and would include a financial incentive of $125 for their time. Participants were invited to attend any to all of the three focus groups in their preferred language, depending on their availability.

### Data Collection

We conducted a total of six focus groups (three each for Spanish- and Cantonese-preferring participants) from February to JuneJune of 2024. Focus groups were held within a conference room in an urban, academic-affiliated county hospital on the same campus as where they were receiving back pain care. Only participants and researchers were present. Each session lasted approximately 90 minutes and was facilitated by native bilingual, bicultural research team members (AGC – native Spanish speaker, PhD, Leadership and Community Engagement specialist and LCF - native Cantonese speaker, MSW, MPH, consultant) with more than 20 years of experience in community outreach and prior experience in medical research. Semi-structured guides were developed as advised by the research team members (PZ, EHD, JY, TB). These guides were initially developed in English and were adapted as culturally and linguistically appropriate from feedback from the bilingual, bicultural focus groups facilitators. The guides were translated into Spanish and Cantonese and backtranslated by the bilingual, bicultural members of the team to check for accuracy (see Appendix 1-3 for focus group guides). Focus group facilitators did not know participants before study commencement and disclosed prior to focus groups their reasons for working on this research study.

Focus groups followed a sequential structure. Earlier sessions (focus groups 1 and 2) aimed to identify patient priorities and barriers to self-management, including probing on participants’ prior experiences with cLBP care and barriers to self-management. The final session (focus group 3) focused on defining patient preferences for the format and content of self-management support materials, including probing on participant suggestions for improving educational resources (Figure 1).

**Figure 1.**
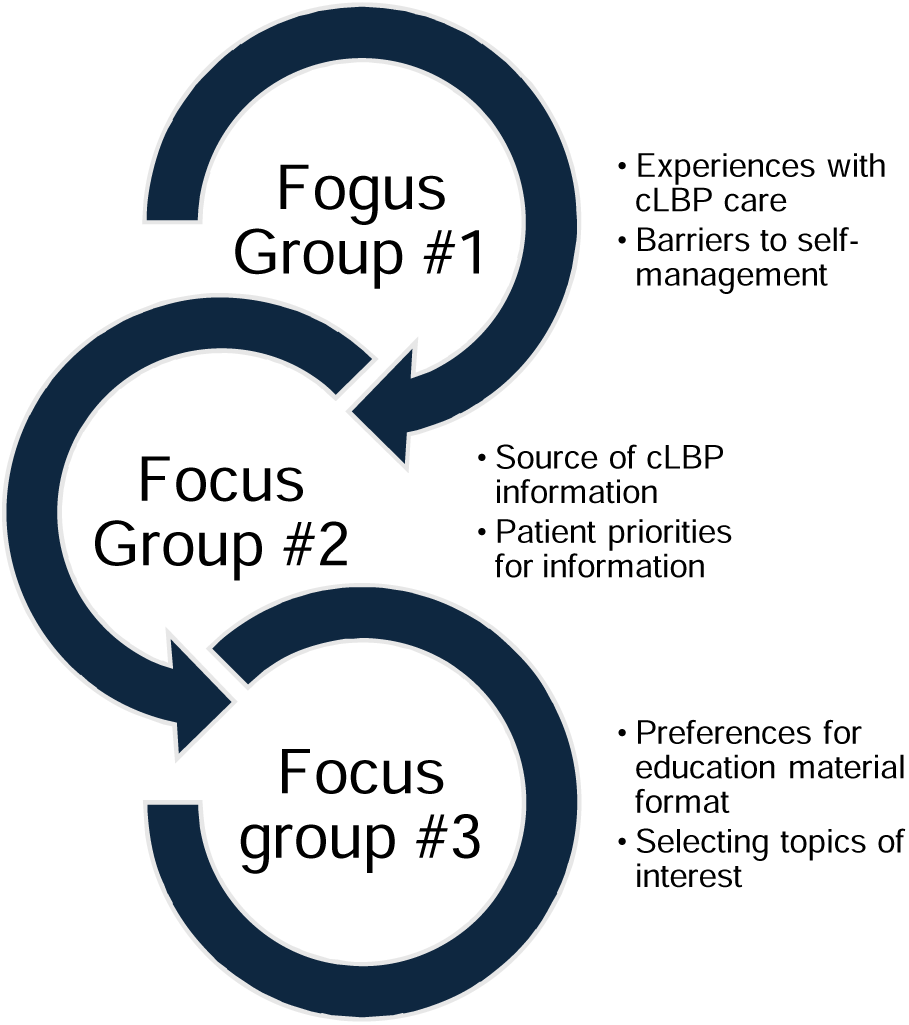
Overview of sequential focus groups.

All focus groups were audio recorded, transcribed verbatim, and translated into English by the company, *transcriptionwing*. Translations underwent a back-translation process by bilingual and bicultural study team members (AK – native Cantonese speaker, AGC, LCF), including the bilingual and bicultural focus group facilitators, to ensure accuracy and cultural relevance. Transcripts were reviewed by the focus group leads for accuracy and conceptual equivalence. Transcripts were not returned to participants for comments. Field notes were made after focus groups.

Participants self-completed a brief survey that included demographic information, household income, education level, language preferences, the P assess pain severity,^36^ numerical pain rating scale,^37^ a 10-point likert scale measure of trust in their healthcare providers.

### Data Analysis

We employed an inductive approach to analyze the focus group data. Two trained qualitative researchers (EHD and JY) independently reviewed the transcripts. Codes were developed to focus on identifying participant priorities for self-management support, barriers to care, and design preferences for educational materials. The two researchers compared their codes and reconciled differences through discussion. Key themes were identified through iterative analysis after the data was collected, with emerging concepts further refined in collaboration with additional study team members (PZ). Microsoft Excel was used.

In our analyses, we highlight differences, when identified, between the Spanish- and Cantonese-preferring focus groups, such as differing views on pain management efficacy and experiences of resource availability. Participants did not provide feedback on the findings.

## Results

### Participant Demographics

A total of 27 unique patients enrolled in the study; 15 participated in focus groups. Two patients withdrew after initial consent and did not provide reason. Twelve patients enrolled but did not participate due to scheduling conflicts. Of the focus group participants, 8 (53%) identified as Latinx and preferred to speak Spanish, and 7 (47%) identified as Asian and preferred to speak Cantonese. The mean age of participants in the Spanish-speaking group was 55.1 (SD 12.2) years, while the mean age in the Cantonese-speaking group was 69.0 (SD 2.0) years. Participants reported moderate to severe pain levels, with an average score of 6.82 on the numerical rating scale of pain (NRS) (SD 2.49). Participants also expressed high levels of trust in their healthcare providers, with 71% of Spanish-speaking and 78% of Cantonese-speaking participants reporting 10/10 “complete trust” in their healthcare providers. See Table 1 for full details.

**Table 1.** Demographics of Focus Group Participants.

|  | Spanish-preferring |  |  |  | Cantonese-preferring |  |  |  |
| --- | --- | --- | --- | --- | --- | --- | --- | --- |
|  | Total | FG#1 | FG#2 | FG#3 | Total | FG#1 | FG#2 | FG#3 |
| # of participants | 8 | 4 | 4 | 4 | 7 | 5 | 7 | 5 |
| Age (mean; SD) | 55.1; 12.24 | 49.0; 14.02 | 55.3; 7.27 | 58.0; 11.11 | 67.3; 4.82 | 69.0; 2/- | 67.3; 4.82 | 66.6; 5.55 |
| #/% Female | 5 (62.5%) | 2 (50.0%) | 2 (50.0%) | 4 (100.0%) | 4 (57.1%) | 3 (60.0%) | 4 (57.1%) | 3 (60.0%) |
| \$50,000 household income or less (#/%) | 6 (75.0%)* | 3 (75.0%)* | 2 (50.0%)* | 2 (50.0%)* | 7 (100.0%) | 5 (100.0%) | 7 (100.0%) | 5 (100.0%) |
| High school graduate or less (#/%) | 5 (62.5%) | 2 (50.0%) | 2 (50.0%) | 4 (100.0%) | 7 (100.0%) | 5 (100.0%) | 7 (100.0%) | 5 (100.0%) |
| Financial difficulties (#/%)<sup>1</sup> | 3 (37.5%) | 2 (50.0%) | 2 (50.0%) | 0 (0.0%) | 6 (85.7%) | 4 (80.0%) | 6 (85.7%) | 5 (100.0%) |
| NRS (mean; SD) | 6.9 (2.4) | 6.3 (2.9) | 7.8 (2.6) | 7.8 (1.7) | 6.4 (1.8) | 6.2 (2.2) | 6.4 (1.8) | 6.0 (1.0) |
| “Complete” trust in healthcare providers (#/%)<sup>2</sup> | 6 (75.0%) | 3 (75.0%) | 2 (50.0%) | 3 (75.0%) | 6 (85.7%) | 5 (100.0%) | 6 (85.7%) | 4 (80.0%) |

1. Answered "How hard is it for you (and your family) to pay for the very basics like food, medical care, and heating?" Answering “very hard,” “hard,” or “somewhat hard” was considered positive for financial difficulties, versus “not hard at all” 2. Answered “How much do you trust your health care provider(s) at this clinic?” Answering 10/10, “Completely,” on a 10 point likely scale was considered “complete trust”
\* All others answered “don’t know”

### Themes and key findings

Four main themes emerged from the focus groups related to patient priorities for self-management support and preferences for educational materials. These themes were consistent across both Spanish- and Cantonese-preferring participants, although specific concerns varied slightly between the two groups (Table 2).

**Table 2.**
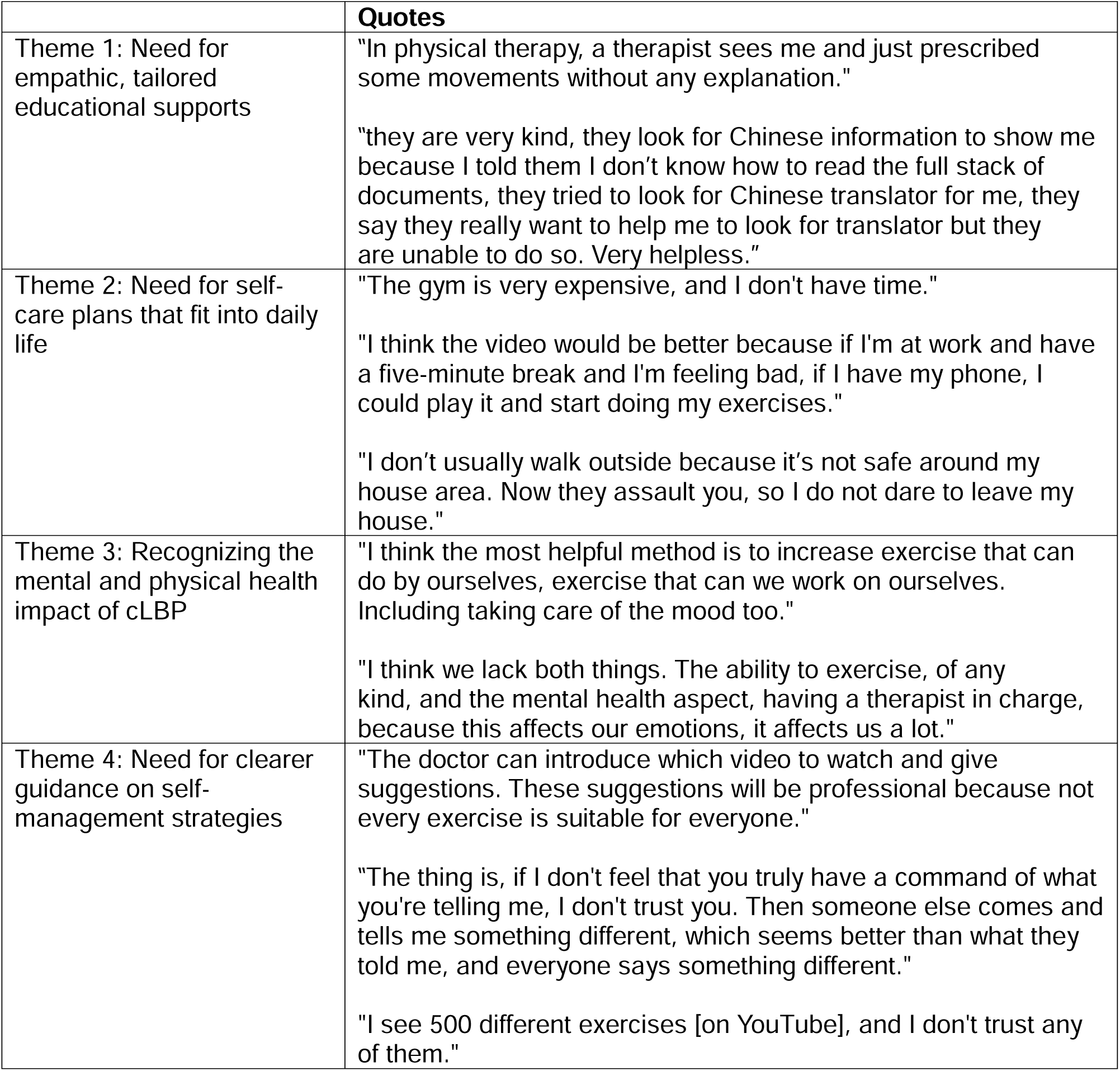
Major themes identified through focus groups.

### Theme 1: Need for empathic, tailored educational supports

Participants across both Spanish- and Cantonese language focus groups expressed dissatisfaction with current cLBP care plans, often describing them as unclear, ineffective, or not aligned with their personal experiences. Participants reported a lack of understanding about the causes of their pain and how treatments such as physical therapy or exercise could help.

Cantonese-preferring participants emphasized the “incurable” nature of their pain and voiced concern that self-management strategies like exercise or physical therapy would be ineffective. Some participants conflated prescription medication with self-care and did not view non-pharmacologic interventions as helpful. As one Cantonese-preferring participant noted: “It [physical therapy] was not helpful. Cannot cure my nerve pain. Regardless of what is done, my nerve was damaged. It is incurable.”

Spanish-preferring participants highlighted the need for better-quality educational materials, with many expressing frustration that information provided by clinicians was either confusing or not helpful. There was a strong desire for clear explanations of their diagnoses and why particular treatments were recommended. As a Spanish-preferring participant noted: “My primary care doctor referred me to physical therapy. In physical therapy, a therapist sees me and just prescribed some movements without any explanation.”

Participants in both focus groups strongly preferred self-management materials that were visually engaging, easy to understand, and available in their preferred language. Short videos were the most requested format, with participants emphasizing the need for clear, simple instructions that could be accessed via smartphone, which may relate to the high levels of digital access that participants noted on surveys. Spanish-speaking participants favored shorter, mobile-friendly videos, while Cantonese-speaking participants preferred written materials with accompanying visual aids, such as diagrams or pictures. Both groups highlighted the need for material that is culturally relevant and uses empathetic communication andand a recognizable “face” or trusted authority to deliver the information.

### Theme 2: Need for self-care plans that fit into daily life

Participants, particularly those in the Spanish-speaking groups, emphasized the difficulty of incorporating self-care practices into their daily routines, especially given their social and economic circumstances. Many participants reported working multiple jobs, lacking access to exercise facilities, or facing financial constraints that limited their ability to engage in recommended activities like yoga or physical therapy. One Spanish-preferring participant noted: “The gym is very expensive, and I don’t have time.”

Cantonese-preferring participants also reported challenges in accessing safe environments for exercise due to fear of being attacked in their communities, reflecting broader concerns about personal safety. One Cantonese-preferring participant noted: “I don’t usually walk outside because it’s not safe around my house area. Now they assault you, so I do not dare to leave my house.”

Both groups expressed a preference for self-management resources that were easy to follow and accessible at any time, such as short videos or simple written guides. Spanish-speaking participants favored videos that could be accessed via smartphone during brief breaks, while Cantonese-speaking participants requested written materials with clear visuals to help with understanding.

### Theme 3: Recognizing the mental and physical health impact of cLBP

Both Spanish- and Cantonese-preferring participants discussed the profound impact of cLBP on their mental health. Feelings of isolation were common, with participants expressing a desire for their care teams to facilitate connections with others experiencing similar challenges. Group visits, support groups, and community-based exercise programs were suggested as potential solutions to reduce isolation and provide mutual support. A Spanish-preferring participant noted: “Yes, it would be good to have courses or workshops, that you can attend. Maybe one or two times a week, that would be something I would like.” Cantonese-preferring participants also recognized the need for more continuity in their care, noting that turnover among clinicians could feel isolating and made it difficult to build trust and follow through on self-management plans.

### Theme 4: Need for clearer guidance on self-management strategies

Participants across both groups expressed difficulty in determining which materials or online resources were trustworthy. Many noted the overwhelming amount of conflicting information available online and stressed the importance of receiving guidance directly from clinicians. As a Cantonese-speaking participant noted: “The doctor can introduce which video to watch and give suggestions. These suggestions will be professional because not every exercise is suitable for everyone.” Spanish-speaking participants also noted feeling unsure about which exercises were appropriate for their specific condition: “I see 500 different exercises [on YouTube], and I don’t trust any of them.”

Despite reporting high levels of trust in their healthcare providers on surveys, both groups commented on skepticism about self-management strategies and confusion about how to approach their care given differences in opinions from different sources. One Spanish-speaking participant voiced, “The thing is, if I don’t feel that you truly have a command of what you’re telling me, I don’t trust you. Then someone else comes and tells me something different, which seems better than what they told me, and everyone says something different.” Both groups called for clearer explanations of the causes of their pain, what self-management entails, and what they could realistically expect from different treatment options. They expressed a need for resources that provide step-by-step instructions, as well as visual aids like videos and diagrams to help them better understand their care plans.

## Discussion

This study aimed to explore the self-management needs and priorities of Spanish- and Cantonese-preferring patients with chronic low back pain (cLBP) at an urban county hospital. By using a HCD approach, we were able to identify key areas where our study populations face barriers to care and where improvements in self-management resources could be made. The findings reveal that patients faced significant challenges in accessing appropriate resources, understanding care plans, and integrating self-management practices into their daily lives. Our results suggest the need for more culturally tailored, accessible, and comprehensive self-management support systems, and build upon prior research on the need for accessible and patient-centered cLBP self-management support.^38–40^

One of the major findings of this study was the pervasive sense of skepticism and confusion regarding current cLBP treatment plans, particularly among Cantonese-preferring participants. Despite reporting high levels of trust in their healthcare providers, participants expressed frustration with care teams who failed to explain the underlying causes of their pain or how prescribed treatments would help alleviate it. This is a common problem with cLBP care as 80 to 90% of cLBP is considered nonspecific – without specific identifiable causes.^41^ While in other populations, patients report existing acceptance that self-care is an integral part of cLBP management,^42^ in our participants, the lack of clarity contributed to a perception that self-care, particularly physical therapy, was ineffective, leaving patients feeling that their pain was incurable. Studies are needed to elucidate whether non-English preferring patients have differing perception of self-management and effort should be dedicated to developing tailored information for these populations.

Participants from both groups specifically requested additional supports to help them navigate the overwhelming amount of health information available online. To address these concerns, participants called for clear guidance on what self-management entails, including the role of physical activity, the expected outcomes of different treatments, and how to safely perform exercises. Education on cLBP causes and self-management strategy is widely supported by multiple professional society practice guidelines.^43,44^ A review of 41 studies on the health information needs of low back pain patients showed that patients want information about the causes of their condition, its prognosis, and self-management strategies.^45^ There was a consistent demand for clear, consistent information presented in understandable language. Our focus group participants echoed these sentiments. Multiple focus group participants referenced looking online for available health information. Prior research found that most cLBP related information online are classified as advertising with variable quality of information.^46^ . As a result, both groups stressed the need for materials that have been “verified” by trusted health professionals.

The language barrier experienced by non-English preferring cLBP patients creates unique needs and may contribute to their cLBP as well as difficulties accessing self-management care. Studies have shown that in general, non-English speakers have worse outcomes than those who are English-proficient.^47^ Studies have highlighted specific strategies to help patients overcome linguistic and cultural barriers to seeking care. For example, among Spanish-speaking populations in the United States, the use of community health workers—known as *promotores*—and multimedia tools like photo comics (*fotonovelas*) that are culturally and linguistically adapted have been highlighted as effective approaches.^48^ Analysis of efforts to develop cancer screening educational materials for Cantonese speakers emphasized the importance of incorporating sociocultural values and health beliefs.^49^ Similarly, our participants stressed the importance of having material presented in their native languages in clear and understandable terms.

Our findings also highlight potential ways social determinants of health (SDOH) may make it difficult for non-English preferring participants to pursue self-management. Spanish-preferring participants specifically reported dissatisfaction with self-care recommendations, noting that the recommendations were difficult to implement given their socioeconomic constraints. Participants, many of whom were working multiple jobs, also highlighted the challenge of finding the time and resources to follow prescribed self-management activities. Additionally, they raised concerns about how mental health and competing life demands, such as financial and familial responsibilities, further hindered their ability to prioritize self-care. While walking programs have been found to be a low cost, effective, and “easily accessible” form of intervention for cLBP,^50^ Cantonese-preferring participants commented on concerns over safety to be able to walk outside for exercise. Socioeconomic status has been strongly associated with cLBP outcome^51^ and our studies highlight how the SDOH of our participants may impact their ability to adhere to self-management. Educational and self-care information should account for the target audience’s specific socioeconomic constraints.

Another major theme was the recognition of how cLBP affected participants’ mental health and contributed to feelings of isolation. Feelings of isolation may be particularly notable in populations with language barriers, which may make it more difficult to find social connections.^52^ Both groups expressed a desire for their care team to serve as a point of connection, helping them link with other patients experiencing similar challenges. Group-based care, including workshops, exercise classes, and support groups, was suggested as a means of reducing isolation and promoting mental and emotional well-being. The importance of addressing mental health as a component of self-management was particularly salient for Spanish-speaking participants, who called for greater access to mental health resources and social support services. Indeed, group based cognitive behavioral treatment has been shown to improve back pain outcomes and be cost effective in other countries.^53^ Support groups without clinician involvement have also been found to be helpful in increasing functional ability and activity and decreasing need for healthcare seeking.^54^ These types of interventions are not widely available in our practice setting. Healthcare systems should explore ways to collaborate with community organizations to create safe, culturally relevant spaces for self-management, such as group-based therapy or peer support programs.

This study is among the first to explore the specific self-management needs of Spanish- and Cantonese-preferring patients with cLBP, populations that have historically been underserved in the healthcare system. Our findings underscore the need for tailored self-management resources that not only address linguistic barriers but also consider cultural, social, and economic factors. Future interventions aimed at reducing cLBP disparities should prioritize the development of culturally appropriate educational materials, including short videos, podcasts, and written guides that are accessible, easy to understand, and actionable.

### Limitations

This study has a number of limitations. First, this study was done at a single urban academic hospital with a convenience sample of patients who prefer to speak Spanish or Cantonese. The views expressed may not reflect those of patients in other settings. Second, the study is subject to selection and social desirability bias. From surveys, we know that there were demographic differences between participants in the Spanish versus Cantonese language focus groups. Spanish-preferring participants were overall younger and active in the work force; Cantonese-preferring participants tended to be older and out of the workforce. These differences may reflect some of the differences in perspectives between the two sets of focus groups that could have been more related to age and life experience, not true differences related to language or culture. Patients from our study setting who declined to participate may have perspectives not captured by the study population. However, across both focus groups, participants expressed similar perspectives, decreasing the likelihood of selection bias. Participants also highlighted a range of challenges of the current self-management resources and care they were being provided, decreasing the likelihood of social desirability bias. As a qualitative study, the goal was to capture insights from a specific patient population in our study setting to inform development of tailored self-management educational material.^55^ Further refinement of self-management materials should strive to capture input for a larger patient population.

## Conclusion

In conclusion, this study highlights the significant barriers that Spanish- and Cantonese-preferring patients face when attempting to self-manage their cLBP. The findings point to a pressing need for culturally sensitive, accessible, and practical self-management support systems that address both the physical and mental health challenges of living with cLBP. Future work should focus on developing and evaluating interventions that can reduce cLBP disparities by enhancing patient education, fostering social connections, and providing clear, reliable, and motivating self-management resources.

## Data Availability

All data produced in the present study are available upon reasonable request to the authors

## Notes

Research reported in this publication was supported by the National Institute of Arthritis and Musculoskeletal and Skin Diseases of the National Institutes of Health under Award Number U19AR076737. The content is solely the responsibility of the authors and does not necessarily represent the official views of the National Institutes of Health. The Core Center of Patient-centric, Mechanistic Phenotyping in Chronic Low Back (REACH) investigators include the following University of California, San Francisco (unless noted otherwise) personnel in alphabetical order:

### Competing Interest Statement

All authors have completed the ICMJE uniform disclosure form at www.icmje.org/coi_disclosure.pdf and declare: no support from any organization for the submitted work; JL owns shares of Bioniks, LLC (a company commercializing a depth camera system similar to that being used by the REACH Physical Function and Biomechanics Core), is an Officer for Bioniks and serves on the Board of Directors; JL owns shares of Aclarion and is a consultant for Aclarion (a company which has developed and is marketing the magnetic resonance spectroscopy (MRS) diagnostic tool that is being used for the REACH comeBACK patient cohort); CO owns options of Aclarion; and JB owns options of Bioniks.

### Funding Statement

Research reported in this publication was supported by the National Institute Of Arthritis And Musculoskeletal And Skin Diseases of the National Institutes of Health under Award Number U19AR076737. The content is solely the responsibility of the authors and does not necessarily represent the official views of the National Institutes of Health.

### Author Declarations

IRB approval has been obtained from the University of California, San Francisco

### Summary of Updates

Updated manuscript with more data.

## References

1. James SL, Abate D, Abate KH, et al. Global, regional, and national incidence, prevalence, and years lived with disability for 354 diseases and injuries for 195 countries and territories, 1990–2017: a systematic analysis for the Global Burden of Disease Study 2017. The Lancet 2018;392(10159):1789–1858. DOI: 10.1016/s0140-6736(18)32279-7.

2. Collaborators USBoD, Mokdad AH, Ballestros K, et al. The State of US Health, 1990-2016: Burden of Diseases, Injuries, and Risk Factors Among US States. JAMA 2018;319(14):1444–1472. DOI: 10.1001/jama.2018.0158.

3. Dutmer AL, Schiphorst Preuper HR, Soer R, et al. Personal and Societal Impact of Low Back Pain: The Groningen Spine Cohort. Spine (Phila Pa 1976) 2019;44(24):E1443–E1451. DOI: 10.1097/BRS.0000000000003174.

4. Trost Z, Sturgeon J, Guck A, et al. Examining Injustice Appraisals in a Racially Diverse Sample of Individuals With Chronic Low Back Pain. J Pain 2019;20(1):83–96. DOI: 10.1016/j.jpain.2018.08.005.

5. Chen Y, Campbell P, Strauss VY, Foster NE, Jordan KP, Dunn KM. Trajectories and predictors of the long-term course of low back pain: cohort study with 5-year follow-up. Pain 2018;159(2):252–260. DOI: 10.1097/j.pain.0000000000001097.

6. Batley S, Aartun E, Boyle E, Hartvigsen J, Stern PJ, Hestbaek L. The association between psychological and social factors and spinal pain in adolescents. European journal of pediatrics 2019;178(3):275–286. DOI: 10.1007/s00431-018-3291-y.

7. Anderson KO, Green CR, Payne R. Racial and ethnic disparities in pain: causes and consequences of unequal care. J Pain 2009;10(12):1187–204. DOI: 10.1016/j.jpain.2009.10.002.

8. Green CR, Anderson KO, Baker TA, et al. The Unequal Burden of Pain: Confronting Racial and Ethnic Disparities in Pain. Pain medicine (Malden, Mass) 2003;4(3):277–294.

9. Institute of Medicine Committee on Advancing Pain Research, Care aE. Relieving Pain in America: A Blueprint for Transforming Prevention, Care, Education, and Research. . Washington, DC 2011;National Academies Pr.

10. Mutubuki EN, Luitjens MA, Maas ET, et al. Predictive factors of high societal costs among chronic low back pain patients. Eur J Pain 2020;24(2):325–337. DOI: 10.1002/ejp.1488.

11. Hruschak V, Cochran G. Psychosocial predictors in the transition from acute to chronic pain: a systematic review. Psychology, health & medicine 2018;23(10):1151–1167. DOI: 10.1080/13548506.2018.1446097.

12. Henschke N, Lorenz E, Pokora R, Michaleff ZA, Quartey JNA, Oliveira VC. Understanding cultural influences on back pain and back pain research. Best Pract Res Clin Rheumatol 2016;30(6):1037–1049. DOI: 10.1016/j.berh.2017.08.004.

13. Beneciuk JM, Hill JC, Campbell P, et al. Identifying Treatment Effect Modifiers in the STarT Back Trial: A Secondary Analysis. J Pain 2017;18(1):54–65. DOI: 10.1016/j.jpain.2016.10.002.

14. Chibnall JT, Tait RC. Long-term adjustment to work-related low back pain: associations with socio-demographics, claim processes, and post-settlement adjustment. Pain medicine (Malden, Mass) 2009;10(8):1378–88. DOI: 10.1111/j.1526-4637.2009.00738.x.

15. Chibnall JT, Tait RC, Andresen EM, Hadler NM. Race and socioeconomic differences in post-settlement outcomes for African American and Caucasian Workers’ Compensation claimants with low back injuries. Pain 2005;114(3):462–72. DOI: 10.1016/j.pain.2005.01.011.

16. Ikeda T, Sugiyama K, Aida J, et al. Socioeconomic inequalities in low back pain among older people: the JAGES cross-sectional study. International journal for equity in health 2019;18(1):15. DOI: 10.1186/s12939-019-0918-1.

17. Suman A, Bostick GP, Schaafsma FG, Anema JR, Gross DP. Associations between measures of socio-economic status, beliefs about back pain, and exposure to a mass media campaign to improve back beliefs. BMC public health 2017;17(1):504. DOI: 10.1186/s12889-017-4387-4.

18. Tait RC, Chibnall JT, Andresen EM, Hadler NM. Management of occupational back injuries: differences among African Americans and Caucasians. Pain 2004;112(3):389–96. DOI: 10.1016/j.pain.2004.09.027.

19. Mathieu J, Roy K, Robert M, Akeblersane M, Descarreaux M, Marchand AA. Sociodemographic determinants of health inequities in low back pain: a narrative review. Front Public Health 2024;12:1392074. (In eng). DOI: 10.3389/fpubh.2024.1392074.

20. Al Shamsi H, Almutairi AG, Al Mashrafi S, Al Kalbani T. Implications of Language Barriers for Healthcare: A Systematic Review. Oman Med J 2020;35(2):e122. (In eng). DOI: 10.5001/omj.2020.40.

21. Soled D. Language and Cultural Discordance: Barriers to Improved Patient Care and Understanding. J Patient Exp 2020;7(6):830–832. (In eng). DOI: 10.1177/2374373520942398.

22. Foiles Sifuentes AM, Robledo Cornejo M, Li NC, Castaneda-Avila MA, Tjia J, Lapane KL. The Role of Limited English Proficiency and Access to Health Insurance and Health Care in the Affordable Care Act Era. Health Equity 2020;4(1):509–517. (In eng). DOI: 10.1089/heq.2020.0057.

23. Villaire M, Mayer G. Low Health Literacy: The Impact on Chronic Illness Management. Professional Case Management July-August 2007;12(4). DOI: 10.1097/01.PCAMA.0000282907.98166.93.

24. Woloshin S, Bickell NA, Schwartz LM, Gany F, Welch HG. Language Barriers in Medicine in the United States. JAMA 1995/03/01;273(9). DOI: 10.1001/jama.1995.03520330054037.

25. Bhondoekhan F, Marshall BDL, Shireman TI, Trivedi AN, Merlin JS, Moyo P. Racial and Ethnic Differences in Receipt of Nonpharmacologic Care for Chronic Low Back Pain Among Medicare Beneficiaries With OUD. JAMA Network Open 2023;6(9):e2333251–e2333251. DOI: 10.1001/jamanetworkopen.2023.33251.

26. Du S, Hu L, Dong J, et al. Self-management program for chronic low back pain: A systematic review and meta-analysis. Patient Educ Couns 2017;100(1):37–49. (In eng). DOI: 10.1016/j.pec.2016.07.029.

27. Zhou T, Salman D, McGregor AH. What do we mean by ’self-management’ for chronic low back pain? A narrative review. Eur Spine J 2023;32(12):4377–4389. (In eng). DOI: 10.1007/s00586-023-07900-4.

28. Mauck MC, Aylward AF, Barton CE, et al. Evidence-based interventions to treat chronic low back pain: treatment selection for a personalized medicine approach. Pain Rep 2022;7(5):e1019. (In eng). DOI: 10.1097/pr9.0000000000001019.

29. Riffin C, Pillemer K, Reid MC, LlJckenhoff CE. Decision Support Preferences Among Hispanic and Non-Hispanic White Older Adults With Chronic Musculoskeletal Pain. The Journals of Gerontology: Series B 2016/09/01;71(5). DOI: 10.1093/geronb/gbv071.

30. Alamam DM, Leaver A, Alsobayel HI, Moloney N, Lin J, Mackey MG. Low Back Pain– Related Disability Is Associated with Pain-Related Beliefs Across Divergent Non– English-Speaking Populations: Systematic Review and Meta-Analysis. Pain Medicine 2021/12/11;22(12). DOI: 10.1093/pm/pnaa430.

31. Escobedo LE, Cervantes L, Havranek E, Escobedo LE, Cervantes L, Havranek E. Barriers in Healthcare for Latinx Patients with Limited English Proficiency—a Narrative Review. Journal of General Internal Medicine 2023 38:5 2023-01-31;38(5). DOI: 10.1007/s11606-022-07995-3.

32. Levander XA, VanDerSchaaf H, Barragán VG, et al. The Role of Human-Centered Design in Healthcare Innovation: a Digital Health Equity Case Study. J Gen Intern Med 2024;39(4):690–695. (In eng). DOI: 10.1007/s11606-023-08500-0.

33. Johnston W, Keogh A, Dickson J, et al. Human-Centered Design of a Digital Health Tool to Promote Effective Self-care in Patients With Heart Failure: Mixed Methods Study. JMIR Form Res 2022;6(5):e34257. (In eng). DOI: 10.2196/34257.

34. Chen YP, Woodward J, Shankar MN, et al. MyTrack+: Human-centered design of an mHealth app to support long-term weight loss maintenance. Front Digit Health 2024;6:1334058. (In eng). DOI: 10.3389/fdgth.2024.1334058.

35. Deyo RA, Dworkin SF, Amtmann D, et al. Report of the NIH Task Force on research standards for chronic low back pain. J Pain 2014;15(6):569–85. DOI: 10.1016/j.jpain.2014.03.005.

36. Krebs EE, Lorenz KA, Bair MJ, et al. Development and initial validation of the PEG, a three-item scale assessing pain intensity and interference. J Gen Intern Med 2009;24(6):733–8. (In eng). DOI: 10.1007/s11606-009-0981-1.

37. Childs JD, Piva SR, Fritz JM. Responsiveness of the Numeric Pain Rating Scale in Patients with Low Back Pain. Spine 2005;30(11).

38. Kongsted A, Ris I, Kjaer P, Hartvigsen J. Self-management at the core of back pain care: 10 key points for clinicians. Braz J Phys Ther 2021;25(4):396–406. (In eng). DOI: 10.1016/j.bjpt.2021.05.002.

39. O’Hagan ET, Cashin AG, Traeger AC, McAuley JH. Person-centred education and advice for people with low back pain: Making the best of what we know. Braz J Phys Ther 2023;27(1):100478. (In eng). DOI: 10.1016/j.bjpt.2022.100478.

40. Zhou T, Salman D, McGregor AH. What do we mean by ‘self-management’ for chronic low back pain? A narrative review. European Spine Journal 2023;32(12):4377–4389. DOI: 10.1007/s00586-023-07900-4.

41. Chiarotto A, Koes BW. Nonspecific Low Back Pain. New England Journal of Medicine 2022-05-05;386(18). DOI: 10.1056/NEJMcp2032396.

42. May S. Patients’ attitudes and beliefs about back pain and its management after physiotherapy for low back pain. Physiotherapy Research International 2007/09/01;12(3). DOI: 10.1002/pri.367.

43. Qaseem A, Wilt TJ, McLean RM, Forciea MA. Noninvasive Treatments for Acute, Subacute, and Chronic Low Back Pain: A Clinical Practice Guideline From the American College of Physicians. Annals of Internal Medicine 2017;166(7):514–530. DOI: 10.7326/M16-2367.

44. Bernstein IA, Malik Q, Carville S, Ward S. Low back pain and sciatica: summary of NICE guidance. Bmj 2017;356.

45. Lim YZ, Chou L, Au RT, et al. People with low back pain want clear, consistent and personalised information on prognosis, treatment options and self-management strategies: a systematic review. Journal of Physiotherapy 2019/07/01;65(3). DOI: 10.1016/j.jphys.2019.05.010.

46. Li L, Irvin E, Guzmán J, Bombardier aC. Surfing for Back Pain Patients: The Nature and Quality of Back Pain Information on the Internet. Spine 2001;26(5).

47. Kandula NR, Lauderdale DS, Baker DW. Differences in Self-Reported Health Among Asians, Latinos, and Non-Hispanic Whites: The Role of Language and Nativity. Annals of Epidemiology 2007/03/01;17(3). DOI: 10.1016/j.annepidem.2006.10.005.

48. Hernandez J, Demiranda L, Perisetla P, et al. A systematic review and narrative synthesis of health literacy interventions among Spanish speaking populations in the United States. BMC Public Health 2024 24:1 2024-06-27;24(1). DOI: 10.1186/s12889-024-19166-6.

49. Tu S-P, Yip M-P, Chun A, Choe J, Bastani R, Taylor V. Development of Intervention Materials for Individuals With Limited English Proficiency: Lessons Learned From “Colorectal Cancer Screening in Chinese Americans”. Medical Care September 2008;46(9). DOI: 10.1097/MLR.0b013e31817f0cde.

50. Hendrick P, Te Wake AM, Tikkisetty AS, Wulff L, Yap C, Milosavljevic S. The effectiveness of walking as an intervention for low back pain: a systematic review. European spine journal : official publication of the European Spine Society, the European Spinal Deformity Society, and the European Section of the Cervical Spine Research Society 2010;19(10):1613–1620. (In eng). DOI: 10.1007/s00586-010-1412-z.

51. Karran EL, Grant AR, Moseley GL. Low back pain and the social determinants of health: a systematic review and narrative synthesis. Pain 2020;161(11):2476–2493. (In eng). DOI: 10.1097/j.pain.0000000000001944.

52. Pot A, Keijzer M, Bot KD. The language barrier in migrant aging. International Journal of Bilingual Education and Bilingualism 2020-10-20;23(9). DOI: 10.1080/13670050.2018.1435627.

53. Lamb SE, Hansen Z, Lall R, et al. Group cognitive behavioural treatment for low-back pain in primary care: a randomised controlled trial and cost-effectiveness analysis. The Lancet 2010/03/13;375(9718). DOI: 10.1016/S0140-6736(09)62164-4.

54. Subramaniam V, Stewart MW, Smith JF. The Development and Impact of a Chronic Pain Support Group: A Qualitative and Quantitative Study. Journal of Pain and Symptom Management 1999/05/01;17(5). DOI: 10.1016/S0885-3924(99)00012-3.

55. Leung L. Validity, reliability, and generalizability in qualitative research. J Family Med Prim Care 2015;4(3):324–7. (In eng). DOI: 10.4103/2249-4863.161306.

